# SEX DIFFERENCES IN THE MANAGEMENT OF SUSPECTED STROKE IN A TELESTROKE NETWORK

**DOI:** 10.1101/2023.03.07.23286961

**Authors:** Soledad Pérez-Sánchez, Ana Barragán-Prieto, Juan Bautista Loscertales, Juan Antonio Cabezas Rodríguez, Elena Zapata-Arriaza, Laura Amaya, Andrés Hermosín Gómez, Miguel Ángel Gamero García, Benito Galeano, Javier Fernández, Blanca Pardo Galiana, Ana Domínguez Mayoral, Leire Ainz Gómez, Jose Fernández Navarro, Cristina del Toro, Manuel Medina, Reyes de Torres, Pablo Baena, Francisco Moniche, Roberto Valverde Moyano, Patricia Martínez, Alejandro González, Joan Montaner

## Abstract

**BACKGROUND:** The evolution of ischemic stroke is different according to sex and is one of the main causes of death in women. In the literature, it is not clear if this is due to biological differences or to disparities in medical care. Previous studies have shown that women are less likely to receive acute treatment, and stroke center type is an important predictor of door-to-needle times. We investigated whether women are attended in a similar way to men in the telestroke network with specialized stroke physicians.

**METHODS:** A prospective registry of ischemic strokes recorded in the centralized Andalusian telestroke network (CATI) was analyzed, focusing on sex differences. Demographic data, clinical characteristics (risk factors), neuroimaging data, treatment intervals, and clinical results (the modified Rankin Scale [mRS] score) were collected. Functional outcomes were evaluated using the mRS at 90 days.

**RESULTS:** A total of 3009 suspected stroke patients were attended to in the telestroke network in the last three years, of which 42.74% were women. A total of 69.54% of the men and 63.85% of the women were diagnosed with ischemic stroke (p=0.002). Women were older (p>0.001) and less independent upon arrival (p=0.006) than men. There was no difference in the treatment received or in the treatment intervals between the groups. Importantly, there was no difference in mRS scores at 3 months between sexes. However, at follow-up, women had fewer imaging tests (p = 0.018) and fewer outpatient visits (p<0.001) than men.

**CONCLUSIONS:** No significant difference between men and women has been found in the acute treatment of stroke in a large telestroke network. However, the same is not true for the follow-up and management of patients after the acute phase. This fact supports that strict adherence to protocols, training, and specialization of care and providing equal attention prevents sex differences in stroke treatment and functional outcomes.

## BACKGROUND

Ischemic stroke is a global public health problem and constitutes one of the main causes of disability in adults. The clinical outcome of stroke differs between sexes, representing one of the main causes of death in women globally^1^. Several studies have considered female sex as an independent predictive factor of disability^2^. It has not yet been fully clarified whether this difference is due to sex dimorphism or to treatment and management discrepancies. Women receive a lower quality of care, have less frequent use of medication-based guidelines and are also less likely to receive thrombolysis than men^3^.

Moreover, women are older than men at stroke onset, which might also be related to a worse outcome, since their life expectancy is longer, and stroke rates increase with age^1^. Although previous studies have shown inconsistent risk factors according to sex, it is generally assumed that strokes of cardioembolic etiology due to atrial fibrillation are more frequent in women, while atherothrombotic strokes are more frequent in men^1,4^. The differences in clinical outcomes after reperfusion therapies according to sex remain unclear; age and stroke severity at onset were found to be confounding factors in previous studies^3,5^.

Innovation in health care and new technologies offer a great opportunity to reduce health care disparities, particularly in stroke. Telestroke is defined as telemedicine-enabled stroke consultation that allows patients to be treated in their local community hospital by an on-site physician with remote consultation by a specialist^6^. The use of telestroke is now a class I recommendation by the American Heart Association^7^. It is believed to have contributed to an overall increase in tPA administration and time to thrombolysis addressing geographic disparities in care. In this sense, it remains to be demonstrated whether these networks not only overcome the geographical problem but also inequalities in terms of gender or race. Previous studies have already pointed to this fact; however, they are still insufficient, and not all have shown results^8^.

In this study, we aimed to explore stroke management and the clinical results according to the sex of patients with suspected strokes managed within a telestroke network.

## METHODS

A prospective registry of ischemic strokes recorded in the centralized Andalusian telestroke network (CATI) was analyzed, focusing on sex differences. Andalusia is a region of Spain in southern Europe with an area of 87,597 km2 and a population of 8,381,944 inhabitants. The telestroke network has six mechanical thrombectomy nodes. Each node is composed of one hub or comprehensive stroke center (CSC), one stroke unit (SU), and several telestroke spoke centers (THs). A vascular neurologist coordinates the program and provides consultations in the morning shift. A pool of vascular neurologists from the different stroke centers is on call for the remaining shift. As a result of this system, there are two vascular neurologists dedicated exclusively to providing 24/7 assistance in all teleconsultations for all THs. In addition to recommending the most appropriate treatment for each patient, the neurologist decides whether the patient should be transferred to an SU or CSC based on the patient’s individual needs and the resources available at each center at that time. Picture archiving communication systems (PACS) and medical records are centralized. Owing to the convenience of having the entire population’s medical records already centralized, network requirements are minimal, consisting mainly of a videoconferencing system (a smartphone with a 4G connection to each center and an on-call neurologist) to allow communication between professionals^9^.

The study was approved by the regional ethics committee, PEIBA (Portal de Ética de la Investigación Biomédica de Andalucía) (protocol number: 1818-N-19).

Demographic data, clinical characteristics, type of stroke, and risk factors (including age, diabetes, dyslipidemia, hypertension, smoking, alcohol intake, atrial fibrillation, and personal history of prior stroke/transient ischemic attack and heart disease) were collected for every patient. Neuroimaging evaluation of the current stroke was performed using computed tomography (CT), from which the ASPECTS score, and the occluded vessel were determined.

We collected data regarding treatment intervals. They were measured in each phase of the process (door-to-CATI time, door and CATI-to-brain imaging time, door and CATI-to-needle time, door-origin and door-to-puncture femoral time, door-origin, and puncture-to-recanalization time).

Stroke severity was measured using the National Institute of Health Stroke Scale (NIHSS) performed by neurologists by videoconference. Regarding follow-up, etiology, neuroimaging, and follow-up visits were recorded. Functional outcomes were evaluated with the modified Rankin Scale (mRS) at 90 days.

### Statistical analysis

Two-sample t tests or chi-square tests were used to compare the clinical characteristics between men and women. Candidate covariates included were age, sex, age-by-sex interaction, NIHSS score at admission, diabetes mellitus, hypertension, atrial fibrillation, heart failure, coronary artery disease, atrial fibrillation, high blood pressure, and prior stroke.

Differences between women and men in stroke care in the acute phase (treatment interval times), follow-up and stroke outcomes (NIHSS score at discharge and mRS at 90 days) were explored using the chi-square test.

All statistical analyses were performed using STATA version 17 (STATA Corp., College Station, TX). All reported p values were based on 2-tailed analyses; statistical significance was set at p <0.05.

### Data availability

The present study is compliant with the journal’s data availability standards, and any data not provided in the article may be shared by request of other qualified investigators. The authors have full access to all the data in the study and take responsibility for its integrity and data analysis.

## RESULTS

A total of 3009 suspected stroke patients were attended to within CATI from 2019 to 2021, of whom 1286 (42.74%) were women. Women were older than men (69.96±15.77 vs. 68.15±13.15 years, p<0.001), and a total of 69.54% of the men and 63.85% of the women had been diagnosed with ischemic stroke (p=0.002). Stroke mimics were diagnosed in a higher proportion in women than men (22.70 vs. 16.34%, p=0.001). The presence of diabetes mellitus, dyslipemia, smoking, alcohol intake, previous stroke or TIA and ischemic heart disease was significantly higher in men. In contrast, atrial fibrillation (31.55 vs. 26.07%, p=0.038) was more frequent in women than in men. Regarding the ischemic stroke etiology, more cardioembolic strokes were found in women than men (39.20 vs. 26.75%, p<0.001). There was no difference by sex in the etiology of patients with hemorrhagic strokes (p=0.781). Complete baseline characteristics according to sex are shown in Table 1. Women also had more severe strokes, with higher NIHSS scores (median 5, IQR 1-10 vs. 4 IQR 1-9, p=0.004).

**Table 1.** Baseline Characteristics

|  | **Women (n=1286)** | **Men (n=1723)** | **P** |
| --- | --- | --- | --- |
| **Age, years (mean, standard deviation (SD))** | 69.96 (15.77) | 68.15 (13.16) | **<0.001** |
| **Medical history, (n,%)** |  |  |  |
| Atrial fibrillation/flutter | 289 (31.56) | 311 (26.08) | **0.005** |
| Diabetes mellitus | 366 (38.73) | 574 (44.91) | **0.004** |
| Hypertension | 829 (75.78) | 1058 (73.42) | 0.178 |
| Hyperlipidemia | 453 (46.70) | 664 (51.0) | **0.043** |
| Smoker | 103 (15.9) | 348 (34.87) | **<0.001** |
| Alcohol intake | 23 (3.79) | 263 (29.48) | **<0.001** |
| Prior stroke/Transient Ischemic Attack | 218 (24.77) | 359 (30.02) | **0.008** |
| Cardiopathy | 152 (17.63) | 266 (23.23) | **0.002** |
| **Clinical characteristics at presentation** |  |  |  |
| National Institute of Health Stroke Scale (NIHSS) score at admission, median (IQR) | 5 (1-10) | 4 (1-9) | **0.004** |
| Prestroke mRS (modified Rankin Scale) score ≤2 (n, %) | 1023 (87.06) | 1403 (90.40) | **0.006** |
| ASPECT≥6 (n, %) | 1683 (97.68) | 1236 (96.11) | **0.013** |
| **Treatment** |  |  |  |
| Thrombolysis | 104 (10.46) | 171 (12.09) | 0.215 |
| Thrombectomy | 137 (13.78) | 164 (11.60) | 0.111 |
| Thrombolysis + Thrombectomy | 83 (8.35) | 120 (8.49) | 0.906 |

Regarding radiological data, a higher percentage of men presented a favorable ASPECT (better than or equal to 6) than women in the initial CT (97.68 vs. 96.11, p=0.013). The main locations of large vessel occlusion were M1 (37.63%) and M2 (23.48%) in women, while in men, they were M1 (25.6%) and ACI (22.47%) (p<0.001). There was no significant difference in reperfusion treatment (24.25% in women vs. 24.91% in men, p=0.699). There was also no difference when the treatment was analyzed separately, either in rtPA (13.90% in women vs. 15.98% in men, p=0.123) or in endovascular treatment (18.76% vs. 17.69%, p=0.456), between groups. Regarding treatment intervals, there were no differences between sexes. Details are shown in Table 2.

**Table 2.** Differences in time intervals between women and men

|  | **Women** | **Men** | **p** |
| --- | --- | --- | --- |
| **Time from symptom onset to arrival at emergency department (ED)**<br>(mean, SD) | 164.39 (389.07) | 187.09 (323.69) | 0.190 |
| **Time from arrival at ED to CATI activation** (mean, SD) | 90.19 (258.02) | 87.73 (299.76) | 0.829 |
| **CATI-to-CT scanner time**<br>(mean, SD) | 59.12 (325.14) | 50.55 (113.05) | 0.408 |
| **Door-to-needle time**<br>(mean, SD) | 89.92 (46.53) | 89.99 (107.26) | 0.993 |
| **Time door-in-out** (mean, SD) | 159.63 (138.18) | 174.46 (199.73) | 0.248 |
| **Door Origin-to-arterial puncture**<br>(mean, SD) | 253.62 (90.63) | 260.12 (126.66) | 0.657 |
| **Puncture-to-reperfusion time**<br>(mean, SD) | 31.41 (32.49) | 44.19 (40.47) | **0.009** |

A total of 90.23% of men and 84.25% of women were admitted to one of the network hospitals (p<0.001), but there was a difference in the service on admission. Men were admitted more frequently to a neurospecialized service (stroke unit, neurology ward or neurointensive care) than women (52.34% vs. 60.40%, p=0.001). Details are shown in Table 3.

**Table 3.** Differences on admission unit between women and men

|  | **Women** | **Men** | **p** |
| --- | --- | --- | --- |
| **Stroke Unit** | 316 (35.59) | 482 (37.60) | 0.339 |
| **Neurology Ward** | 48 (5.41) | 97 (7.57) | **0.047** |
| **Neurointensive Care** | 67 (7.55) | 123 (9.59) | 0.097 |
| **Internal Medicine** | 344 (38.74) | 434 (33.85) | **0.020** |

At follow-up, women had fewer neuroimaging tests than men (69.51 vs. 74.05%, p=0.018). Among patients who had follow-up neuroimaging, CT was performed in a greater number of women (32.61 vs. 26.70%, p=0.011) and MRI in a greater number of men (65.22% vs. 71.02%, p=0.015).

Follow-up visits were conducted for 29.61% of patients with differences by sex (25.93% in women vs. 32.57 in men, p<0.001). The global mortality rate was 14.86% without a difference between women and men (15.37% vs. 14.47%, p=0.508), even after adjustment for confounders (mRS score, age, atrial fibrillation, previous TIA/stroke) (OR 0.81; CI95% 0.61–1.07). There was no significant difference by sex in good functional outcomes (mRS ≤2) at discharge (72.80 vs. 72.91%, p=0.965) and 90 days (71.58 vs. 76.97%, p=0.179) among surviving patients. After adjustment for confounders (age, diabetes, dyslipidemia, and ischemic heart disease), these data were maintained (OR 0.93, CI95% 0.59–1.47).

## DISCUSSION

Women have a higher burden of stroke mortality and disability than men^10^. However, it is not clear whether this is due to purely biological differences in the response to brain ischemia and ischemic risk or to a disparity in health care provision^1^. Andalusia has been called the “Spanish Stroke Belt” on account of its high stroke mortality rate. A telestroke network was created to improve this statistic by reorganizing and improving stroke care in the region. In this way, inequalities between areas have been reduced^11^.

Regarding the acute phase and differences in health care provision between sexes, our data show that there are no important differences in the management of stroke in the acute phase with similar rates of treatments and interval times to treatment. Previous studies have shown that women are less likely to receive endovascular treatment or rtPA than men, even after adjusting for functional status before onset and the time from onset to the hospital door^16,17^. Stroke center type was also an important hospital-level predictor of tPA utilization and times to treatment in previous studies, with differences in patients treated in a rural/smaller hospital and those treated in a metropolitan/larger hospitals^18,19^. Hospitals without specialized stroke care have longer delays, which is pronounced for women^5^. In our study, we found more proximal occlusions in men, mostly of atheromatous origin, and more distal occlusions in women, related to AF in most cases. This may be one of the causes of treatment delays in women due to the difficulty of diagnosis and presentation (stroke mimics are also more frequent in women). However, this aspect has been overcome with telestroke networks and specialized physicians. On the other hand, we found longer times for reperfusion in men, probably related to these proximal occlusions of atheromatous origin that make the procedure more difficult.

Most studies have found that strokes are more severe in women than in men, although age may be a confounding factor. Although severity increases with age, previous studies have shown poor outcomes, even after adjusting for risk factor profile and age^12^. The differences in the risk factor profile found in men and women seem to be one of the main points of interest. In general, most modifiable risk factors are found more frequently in men; however, women with modifiable risk factors have a greater likelihood of developing a stroke^13^. Moreover, atrial fibrillation and ischemic strokes are more frequent in women^14,15^. These profiles are also met in our study.

Although functional outcomes after stroke usually differ according to sex, our study demonstrates that with proper management in the acute phase following established protocols by vascular neurologists, these outcomes can be balanced. Thus, the mRS at three months in our study was similar between both sexes. However, although there are no differences in acute management, there are differences in patient follow-up. In this case, this follow-up is not guided by neurologists and is performed according to the center. Men are admitted to the neurology service more frequently than women. It is known that admission to stroke units and treatment by specialized physicians reduces mortality and dependency and is recommended by the main management guidelines. Even for hemorrhagic strokes with high dependency and subsidiary mortality, admission to the stroke unit is better than admission to intensive care^20^. This different management may influence outcomes, and perhaps, the degree of dependence and mortality could be lower in women if they were treated the same as men.

Prior studies have shown that women are less likely than men to be investigated with standard diagnostic tests and that women admitted to hospitals with stroke are less likely to receive defect-free care^11^. In our study, women had fewer neuroimaging tests after stroke. This fact causes women to have a higher percentage of misdiagnoses. This could be related to admissions to nonspecialized units, internal medicine in most cases, which also results in poor follow-up. Only one-third of patients have any follow-up visits in the months after acute treatment, and most of these patients are men. This may affect our results since we do not have complete data for all patients. However, this indicates the need for specialized physicians with adequate training who can guarantee a complete study of all patients to ensure adequate secondary prevention.

Much effort has been made to reduce gender differences in the acute management of stroke, but there is still a significant gap in subacute management. These differences could be overcome with a comprehensive strategy that addresses the existing issues, including further education of physicians and written protocols, remote support from tertiary care institutions, and implementation of future subacute clinical trials that test rural/urban strategies for stroke care.

